# Perceptions and experiences of different Long COVID community rehabilitation service models from the perspectives of people living with Long COVID and healthcare professionals

**DOI:** 10.1101/2023.08.23.23293876

**Authors:** Edward Duncan, Lyndsay Alexander, Julie Cowie, Alison Love, Jacqui Morris, Rachel Moss, Jane Ormerod, Jenny Preston, Joanna Shim, Emma Stage, Tricia Tooman, Kay Cooper

**Affiliations:** University of Stirling; Robert Gordon University; Long COVID Scotland; University of Dundee; NHS Healthcare Improvement Scotland; Douglas Grant Rehabilitation Centre, Ayrshire Central Hospital

## Abstract

**Objectives:** To explore the perceptions and experiences of barriers and facilitators to accessing Long COVID community rehabilitation.

**Design:** We used a qualitative descriptive [1] design over two rounds of data collection with three participant groups: i) people with experience of rehabilitation for Long COVID (PwLC); ii) NHS staff delivering and/or managing community rehabilitation services (allied health professionals (AHPs)), and iii) NHS staff involved in strategic planning around Long COVID in their health board (Long COVID leads).

**Setting:** Four NHS Scotland territorial health boards.

**Participants:** 51 interviews: eight Long COVID leads (11 interviews); 15 AHPs (25 interviews) and 15 PwLC (15 interviews).

**Results:** Three key themes were identified: i) Accessing care for PwLC, ii) Understanding Long COVID and its management; and iii) Strengths and limitations of existing Long COVID rehabilitation services.

**Conclusions:** Organisational delivery of Long COVID community rehabilitation is complex and presents multiple challenges. In addition, access to Long COVID community rehabilitation can be challenging. When accessed, these services are valued by PwLC but require adequate planning, publicity, and resource. The findings presented here can be used by those developing and delivering services for people with Long COVID.

**Strengths and limitations of this study:**

- This is the first study to explore perceptions and experiences of different community rehabilitation models for Long COVID in the Scottish context
- Perspectives of people with Long COVID, staff delivering services, and staff leading on Long COVID in four geographically diverse Scottish health boards are included
- The strengths and limitations of different Long COVID rehabilitation service model components are highlighted
- The rapidly evolving nature of Long COVID and its management resulted in fewer distinct differences between the four health boards than originally anticipated

## INTRODUCTION

### Background

Long COVID, a condition defined as ongoing COVID-19 symptoms that continue beyond twelve weeks following an initial acute COVID-19 infection, was first described in Spring 2020 [2]. It is estimated that at least 65 million people globally have Long Covid [3]. By January 2023, over 2 million people were estimated to have Long COVID in the United Kingdom (UK), including over 175,000 living in Scotland [4]. Long COVID has broad multiple system presentation and can have profound physical, emotional, social, and financial consequences. Moreover, debilitating symptoms of Long COVID often persist for a year or more [3].

NHS funding to support the diagnosis, treatment, and rehabilitation for Long COVID varies across the UK. Since the autumn of 2020, NHS England have allocated £94 million (£1.66 per capita, based on the 2021 UK Office of National Statistics Census data England’s population was 56,489,800) to be invested in specialist Long COVID clinics in England to complement existing primary, community, and rehabilitation care services [5]. The Welsh Government has allocated £18.3 million since 2020 (£5.88 per capita based on a population of 3,107,500 in 2021) to support the delivery of Long COVID diagnosis, treatment, and rehabilitation within existing services [6]. In September 2021, the Scottish Government announced £10 million (£1.82 per capita based on a population of 5,479,900 in 2021) over 3 years to support Scottish NHS health boards’ response to Long COVID [7]. At the end of May 2022, plans for the first £3 million of that funding were announced.

The National Institute for Health and Care Excellence (NICE) [2] and The World Health Organisation [8] recommend people with Long COVID, following an initial medical assessment to exclude underlying conditions, have access to rehabilitation to aid their recovery. As with other long-term conditions, community rehabilitation for people with Long COVID should be personalised, multidisciplinary, and comprehensive in order to maximise function, quality of life, and participation in society [9 10].

However, the optimal approach to deliver Long COVID community rehabilitation is currently unknown. NICE clinical guidance suggests broad rehabilitation approaches, which can be delivered in isolation or in combination, such as self-management and multidisciplinary rehabilitation [2]. The effectiveness of these approaches is not yet clear and there are further uncertainties about the most appropriate means of organising service delivery, particularly that of multidisciplinary rehabilitation. Furthermore, there are well-documented barriers to accessing healthcare services for PwLC [11–13]. Our 2019 national survey [14] of Scotland’s 14 territorial health boards found that Long COVID community rehabilitation was universally available across Scotland. At the time of the survey, one health board was delivering Long COVID community rehabilitation as a dedicated service, with the remainder integrating rehabilitation within existing services. And yet, there was substantial variation across health boards in the mode of service delivery, given that the means of optimally delivering community rehabilitation to this population was unknown.

To date rehabilitation services for Long COVID have varied internationally with respect to delivery and the healthcare professionals involved [15], including virtual rehabilitation focusing on self-management and education [16], group-based pulmonary rehabilitation [17], tiered multidisciplinary pathways [18], and phased approaches to managing symptoms [19]. Our research group are investigating models of service delivery for Long COVID rehabilitation and will provide recommendations for the effective provision of community rehabilitation for PwLC. As part of our larger study, we are exploring the experiences and perceptions of people receiving rehabilitation for Long COVID and NHS staff (operational and strategic) involved in delivering or managing Long COVID community rehabilitation. The aim of this phase of the study was to explore the perceptions and experiences of barriers and facilitators to Long COVID community rehabilitation as services evolved over a 12-month period.

## METHODS

### Study Design

This paper reports on the first two rounds of data collection of a longitudinal qualitative descriptive [1] study based on 51 interviews with three participant groups; i) people with experience of rehabilitation for Long COVID (PwLC); ii) NHS staff delivering and/or managing community rehabilitation services (allied health professionals (AHPs)), and iii) NHS staff involved in strategic planning around Long COVID in their health board (Long COVID leads). The study is reported in accordance with the Consolidated criteria for reporting qualitative research [COREQ] [20].

### Setting

The study was conducted in four Scottish health boards (HB1-HB4). The boards were selected for their geographical and demographic spread as well as variation in Long COVID rehabilitation service models, summarised in Table 1. As shown in Table 1, all boards adopted some form of multi-disciplinary team (MDT) approach using a blended mode of delivery (face-to-face, telephone, Near Me). Staffing varied; occupational therapy and physiotherapy were most consistent across services, with some including additional professions such as dietetics, speech and language therapy, and psychology. Notably, none of the services included medical staff.

**Table 1:**
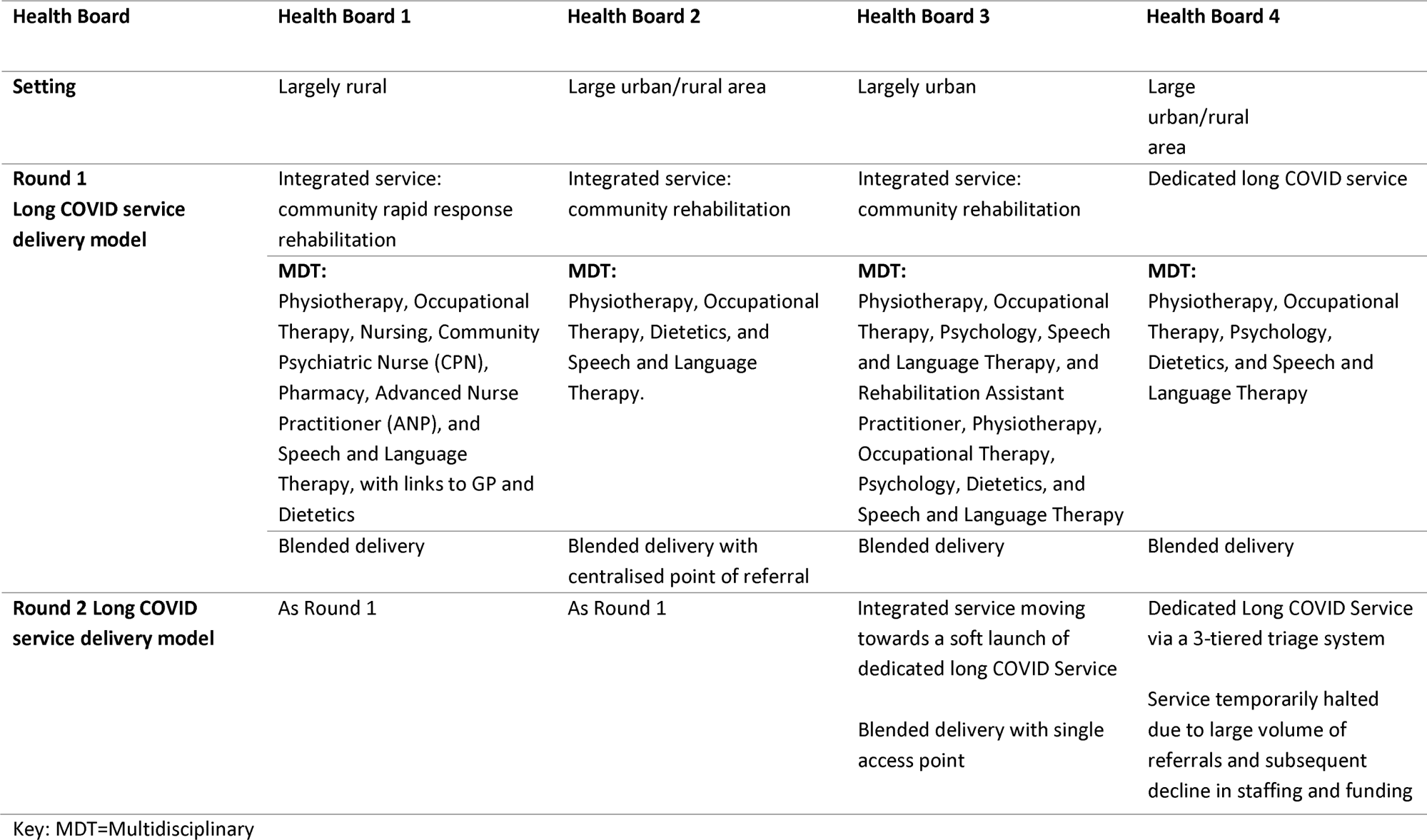
Description of participating health boards.

### Recruitment and Data Collection

Two rounds of data collection took place: Round 1 (November 2021 – April 2022) and Round 2 (June – July 2022). In each round, three participant groups took part: PwLC, AHPs, and Long COVID leads. Long COVID leads were purposively sampled by email invitation or word of mouth, for their role in leading on Long COVID within their health board, and included AHP directors, medical directors, and clinical service leads. AHPs, most often occupational therapists and physiotherapists, were involved in providing Long COVID rehabilitation. Most AHPs acted in a team member role, though a small number were also clinical service leads. AHPs involved in the treatment of PwLC in the study health board areas were purposively recruited by email invitation. The invitation was sent by senior managers on behalf of the study team and explained the reasons for the research. Finally, a convenience sample of PwLC were recruited by their AHP. PwLC were provided with a study pack in printed or electronic format that explained the purpose of the study and volunteers opted into the study by contacting the research team by email, telephone, or leaving their contact details after completing a survey of health-related outcomes as part of the larger study.

Interview topic guides [see Supplemental file 1] were developed by the research team that included people with lived experience of Long COVID. Topic guides were further refined for Round 2 to take account of the temporal nature of the questions being asked. It combined a theory-based approach using the TIDieR intervention description categories [21], and a series of questions exploring facilitators and barriers to delivering and receiving community Long COVID rehabilitation. The interview topic guide was pilot tested with people who had lived experienced of Long Covid and with two AHPs who were not involved in the study prior to implementation. Web-based [Microsoft Teams] or telephone interviews were conducted by five experienced healthcare researchers with master’s or doctoral level qualifications and experience of qualitative research [VB, JC, RM, JS, TT]. All interviewers were female and had experience of qualitative NHS data collection and analysis. Two of the researchers were AHPs and had prior awareness of the challenges of delivering rehabilitation. All interviewers were provided with training and supervision from KC and ED and had no prior relationship with the services or participants. Informed consent was obtained from all participants and recorded verbally prior to each interview. Interviews lasted between 22 and 86 minutes (average 52 minutes). Only the participant and interviewer were present during each interview. Interviews were audio recorded and transcribed intelligently by an external transcription service, where redundant words and/or sounds were removed. Field notes were taken during some interviews. These notes added to team discussions to support understanding of the data as it was collected but did not form part of the official data analysis. Transcripts were not returned to participants for comment or correction.

### Data analysis

Data were analysed using the Framework method, which is commonly used in applied health research and recognised as appropriate for multidisciplinary research teams [22].

For each participant group, at least two researchers from the team familiarised themselves with at least 20% of transcripts, making analytical notes directly on the transcripts. Following discussion between all team members involved in familiarisation, a framework for analysis and interpretation was constructed [23]. Although line-by-line coding is common, it is also possible to develop a framework without engaging in explicit coding [23]; we adopted the latter approach. The ‘analytical framework’ [22] was applied across all interview transcripts using highlighting and the comments function in Microsoft Word, with regular review and discussion within the team. A charting matrix was created in Microsoft Excel that involved summarising broad categories of data from the transcripts, then interpreting the charted data by exploring within and between-cases to derive concepts and themes. These were then grouped into overarching themes. All researchers were involved in interpreting the data during regular team discussions. Participants did not directly provide feedback on the findings. However, the findings were presented in a webinar attended by Long Covid Leads, AHPs and PwLC, several of whom had participated in the study. The webinar attendees endorsed the study findings and reflected that they resonated with their personal experiences.

### Patient and Public Involvement

Two people with lived experience of Long COVID were core members of the research team. They co-developed study materials and interview topic guides and contributed to analysis and interpretation of study findings.

## RESULTS

### Participant characteristics

We recruited eight Long COVID leads, who took part in 11 interviews over the two rounds of data collection: 15 AHPs (25 interviews) and 15 PwLC (15 interviews). See Table 2 for participant details. No participants who consented withdrew from the study.

**Table 2.**
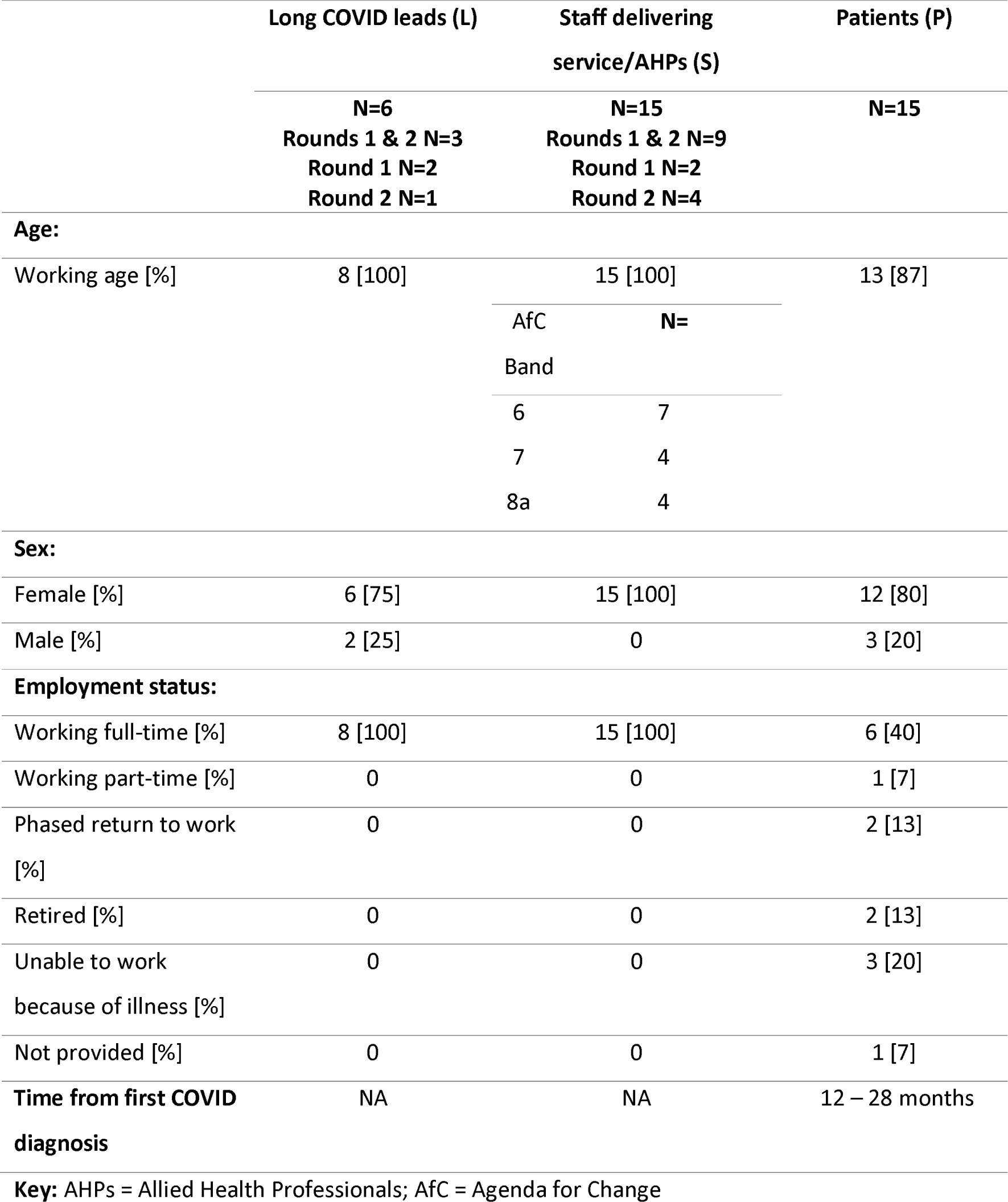
Participant demographics.

### Findings

Framework analysis resulted in three key themes being identified: i) Accessing care for PwLC, ii) Understanding Long COVID and its management; and iii) Strengths and limitations of existing long COVID rehabilitation services. Table 3 illustrates the three themes and the categories of data that contributed to each.

**Table 3:** Key themes and categories of data.

| Key Themes | Sub themes |
| --- | --- |
| **Accessing care for people with Long COVID** | PwLC accessing General Practitioner (GP) services [L; P]<br>GP referring PwLC to community rehabilitation [L; P; S]<br>Awareness of services by GPs & PwLC [S]<br>Signposting [S]<br>Waiting lists & waiting times [P] |
| **Understanding Long COVID and its management** | Long COVID prevalence [L]<br>Long COVID unknowns [L; P]<br>Reluctance to diagnose [P]<br>Tests/investigations [P]<br>Being believed [P]<br>Evolving evidence-base [L; S]<br>Education of PwLC and community rehabilitation staff [S]<br>Information sharing [S]<br>Learning from other conditions [S]<br>Confidence of community rehabilitation staff [S] |
| **Long COVID services** | Current services strengths & limitations [L; P; S]<br>Resourcing services [L; P; S]<br>Need for Long COVID services [L; P; S]<br>Navigating politics [L] |

Key: L = Long Covid leads; P = PwLC; S = AHPs

Participant data was coded and reported as follows: (1) Role of participant: L = Long Covid leads; P = PwLC; S = AHPs; (2) Health Board (HB) number: HBs 1-4; (3) Round of data collection: 1 or 2; and (4) Participant number (e.g. the participant identifier for the first quote: P-3-2-08 is a PwLC from HB3, data was collected in Round 2 and was the 8th PwLC interviewed).

#### I: Accessing care for People with Long COVID

Barriers to accessing Long COVID rehabilitation were identified by all participant groups. In Round 1, patients commonly attributed this to limited availability of face-to-face GP appointments due to COVID-19 restrictions.

> *“You don’t get face-to-face with the GP. So again, when you get a GP, it’s always a different GP you’re talking to and you try your best, but you get locum, and they’ve all been fantastic. I’m not, again, I wouldn’t criticise them, but it’s exhausting telling the same story every single time.” [P3208]*

Long COVID leads and AHPs observed people’s reluctance to seek help from services widely acknowledged as being under pressure. By Round 2, they felt that the perception of GPs being closed, and services overwhelmed, needed to be questioned, and called for greater public awareness of Long COVID rehabilitation services and how to access them.

> *“[We] need to get past the public’s perception that GPs are shut so they return to their GP and get a referral to their service if required.” [L3102]*

All participant groups were aware of the dedicated Long COVID service in HB4. However, due to the publicity the service had received and the pro-active development of a clear GP referral pathway, demand for this service quickly exceeded capacity. Staff (AHPs and Long COVID leads) expressed concern that promoting a Long COVID service in their own board may result in the same situation. Their concerns were compounded by the widely acknowledged pressure in the healthcare system due to a range of factors including staff redeployment, absence, and reopening of non-essential rehabilitation services that had been closed during previous phases of the pandemic.

> *“They* [HB4 Long COVID service]*were basically inundated. So, I had to wait, about four months or so, something like that for a slot. But it was quicker than I thought, but they were just getting overwhelmed with the workload.” [P4101]*

> *“Yeah. So, it’s [integrated service] not a widely publicised thing, because I don’t know if we could cope as a service with the numbers.” [S1102]*

Patients and staff reported both frustration and understanding around long waiting times for Long COVID rehabilitation:

> *“The referral side of it they probably done all the right things, but the timescales were just probably horrific but understandable. I know that in hindsight now so, but it didn’t take away the frustrations of it.” [P3208]*

> *“Difficult due to capacity. Everywhere so busy and up against it. The pressures on us are really extreme.” [L3101]*

The dedicated service in HB4 was accessed by self-referral and delivered by AHP staff. Self-referral was viewed positively by patients; however, staff reported that the lack of information on self-referrals made appropriate triage of patients challenging. Health boards without self-referral and/or a Long COVID service also expressed concern that people with Long COVID would not be prioritised for rehabilitation within usual community rehabilitation services.

> *“I don’t have self-referral at the moment. And the referrals coming through the GP or through something, some of them are coming through. But I don’t know where all the others are, so, I’m not blocking access, but they are getting stuck in the system, in my system, in that if I don’t have enough information on the referral, they’re just going on to the normal routine waiting list.” [S2207]*

HB4 reported the lack of medical staff as a limitation to their dedicated service and, in keeping with other health boards, reported that the lack of a clear Long COVID pathway was a limitation, particularly as people often presented with complex issues that were unsuitable for immediate rehabilitation.

> *“But it’s those that have the more complex needs that I wouldn’t know where to send them to.” [S2103]*

#### II: Understanding Long COVID and its management

Patients commented on the burdensome nature of managing Long COVID and there being a reluctance among GPs to provide a diagnosis. They noted concerns about documenting Long COVID in healthcare records and a need to rule out other underlying conditions.

> *“It’s quite difficult and just feel like, you know, you have a list as long as your arm when you actually speak to the GP because some things have changed and you know they’re very sympathetic, but very clear and honest that well, we don’t really know, you know? We just don’t know. So, we’ll give you this pill to try and treat this symptom at the moment and, that’s quite hard.’ [P4101]*

The lack of diagnosis was also observed by AHPs and Long COVID leads, who shared a discomfort associated with labelling patients.

> *“What are we shaming people for having a condition? What’s that all about? It’s very bizarre. You get them help. I don’t understand. It’s like invisible illnesses.” [L1202]*

> *“It was really difficult actually to know whether to adopt the term Long COVID ‘cause it’s not a medical term, but we’ve decided in the end it’s probably got wider recognition, now, that’s what the patients are using. But having to be very careful, actually even when you’re writing letters to GPs, you know if they didn’t have confirmed COVID in the beginning, it’s a presumed COVID illness, you know symptoms consistent with Long COVID, you know. It’s a lot of kind of working around the houses I feel.” [S4101]*

Most patients expressed a need to be heard and believed by healthcare professionals. They also felt that there was powerful value in hearing that other people with Long COVID experienced similar symptoms, which provided a helpful process in validating patients’ unmet needs.

> *“It felt to me like it was a bunch of very random symptoms. It didn’t seem connected, and I think probably one of the best things in that first conversation was just hearing that the symptoms I have were very common and they are all related.” [P4102]*

> *“I get the impression that a lot of my symptoms are replicated a lot across, across other people so and I’m getting a lot of assurance on what people’s progress like recovery-wise again its always pinned by keep your expectations to a minimum, like it’s no* [not] *going to be the miracle cure.” [P3208]*

Both AHPs and Long COVID leads spoke of the challenges of understanding Long COVID and its constantly evolving evidence base, as well as the lack of data on prevalence. AHPs also reported a lack of evidence to support best practice for managing Long COVID, which could result in anxiety around patient management.

> *“So, what I was concerned about was what if I give the wrong advice or the wrong sort of exercise prescription, and that I actually cause harm to him by something that I’ve done. And I watched some of the podcasts to sort of understand what we should and shouldn’t be doing, and that is when I started to realize that there is a really strong link with Long COVID and the chronic fatigue syndrome and ME population who have been saying for years and years that, actually, sometimes, graded exercise therapy is really harmful, and you absolutely should not prescribe it.” [S2104]*

To increase understanding of Long COVID, AHPs sought out peer support, online resources, and knowledge from other conditions to determine how to support people with Long COVID.

> *“There’s certainly discussions around what is the best way to approach management of Long COVID and there’s been a few things we can do, though, because nationally there’s a huge focus on physiotherapy management of Long COVID you know, the CSP [Chartered Society of Physiotherapists], I’ve got lots of stuff out there, but we’ve also got the clinicians and the COVID rehab team there that although they’re not seeing patients and more than happy to signpost people to resources and help with a bit of professional advice. I would say from a Community hub, clinician point of view, confidence hasn’t been the main issue. They do seek professional advice on how to manage them.” [S3203]*

#### III: Strengths and limitations of existing Long COVID services

A characteristic of all participating health boards during the study period was the dynamically evolving nature of the services provided. This was partly due to external factors relating to COVID prevalence, the restrictions this placed on service delivery, and financial planning decisions. Two health board areas changed their organisational structures affecting services. One health board (HB4) commenced a limited dedicated Long COVID rehabilitation service, delivered by an occupational therapist and a physiotherapist. The service developed clear pathways for referral that gained considerable publicity, as a result of which, the service was unable to meet demand. This was compounded by staff leaving their post. A considerable waiting list developed. The service ran for 18 months after which there was no further funding to continue its delivery, the service ceased and individuals on the waiting list were distributed to local community rehabilitation teams.

#### Strengths

By Round 2 of data collection, AHPs in all health boards were able to see PwLC face-to-face, conduct home visits, have greater integration into the community, and begin working towards return-to-work packages with appropriate Long COVID patients. All these service features were perceived by clinicians to increase the flexibility of service delivery. Patients had experienced a range of modes of service delivery, largely a blended model of face-to-face individual, group, or online interventions. However, irrespective of the delivery model, most patients had a positive experience of Long COVID rehabilitation, especially the self-management information they received. PwLC reported that validation of their experience was particularly significant.

> *“I’m certainly more educated, already daily life has improved I’m not making myself ill doing things I didn’t know were doing that.” [P3203]*

> *“…the relief to at that point to speak to a professional who was actually acknowledging that this was happening and you know, it wasn’t all in my head. I don’t know there, for months and months and months I think it was common there was just no-one apart from my GP, there was just no of recognition or acknowledgement of what people were going through or anything like that. So, it was just such a relief to speak to somebody who was just genuinely interested and very supportive.” [P4107]*

In the two health boards where dedicated Long COVID rehabilitation teams existed, AHPs reported that the MDT approach worked well. It was perceived to increase communication, enabling staff to develop both informal learning about Long COVID through peer discussions and also to develop their knowledge and skills in working with the emerging Long COVID clinical population through in-service learning sessions. The Long COVID leads in these areas perceived that the services were valued by people receiving their care and support, which was corroborated by the patient data.

> *“It was my GP that referred me to the psychologist then obviously the two of them were interacting so that the [COVID team] were speaking to the psychologist as well and just basically building a care package for me, so it was tailored to what I was needing physically and mentally, so they were quite good actually talking to each other as well, which was I suppose positive.” [P4105]*

AHPs and Long COVID leads from all health boards reported high levels of staff engagement in learning about Long COVID. In the absence of specific evidence-based guidance on Long COVID rehabilitation, AHPs reported drawing on information from a wide variety of sources. Sources included a sparse literature that had been published on rehabilitation, guidance from professional bodies such as the Chartered Society of Physiotherapy and Royal College of Occupational Therapists, and learning from established approaches for conditions with similar presenting features, such as functional neurological disorder, chronic fatigue syndrome, and fibromyalgia.

AHPs and Long COVID leads reported that several staff developed an individual professional interest in the condition, worked to increase their knowledge, and sought help to learn from peers locally, across health boards in Scotland, and more widely across the United Kingdom.

> *“England has been incredibly helpful in giving and sharing lots and…you know… even within the LOCO-RISE study that’s been a really helpful kind of network and relationship to support.” [L3203]*

Long COVID leads in health boards where Long COVID rehabilitation services existed reported that these services enabled increased access to care with appropriate referral routes and adaptable and flexible services. One lead described the merit of a blended model of delivery and believed that telephone triage was a useful method of gaining information about patients’ needs.

> *“Do you know, I think the use of the increased use of digital services has been fantastic. It’s meant that we’ve been able to reach people when we wouldn’t have normally been able to reach them. It has to be a blended model. It has to be that those people that need face-to-face intervention get it at a time when it’s right. But digital advances have really made a big difference for us.” [L1101]*

### Limitations

All participating health boards experienced considerable challenges in delivering Long COVID rehabilitation services. A key challenge was funding, as dedicated funding for Long COVID services had not been allocated by the Scottish Government Long COVID fund at the time these data were collected. The two health boards that had developed dedicated services used existing funding allocations. The short-term nature of this funding in one health board meant that their limited service could not be sustained, and that service ceased during Round 2 of our data collection. This limited investment in developing the structure for Long COVID rehabilitation was highlighted by participants as a considerable limiting factor in service delivery, regardless of whether the health board had developed a dedicated service or not, and the *“historical underinvestment in rehab services” [L2101]* was also highlighted by long COVID leads as a significant limiting factor. Where Long COVID dedicated services had not been developed, services reported that they lacked the staffing capacity to meet the perceived additional service demand.

> *I think we’re struggling even staffing wise with. We can’t staff wards properly. We can’t. You know all these things it’s. How will we staff a Long COVID service? How will we? How? How will we best provide that service? The NHS way has always been to kind of just absorb it into existing services.” [S1202]*

However, where individuals with Long COVID were seen by existing rehabilitation services these services (described henceforth as ‘integrated services’) were observed to receive considerably fewer referrals for Long COVID rehabilitation. This appeared to be due to the reluctance in these health board areas to promote services (as discussed above). AHPs in integrated services also reported that their referral criteria were often not appropriate for people with Long COVID, resulting in referrals being rejected as inappropriate as PwLC often did not meet pre-existing service criteria for community rehabilitation. Fewer Long COVID referrals entering the services led to reduced confidence among AHPs in their ability to appropriately manage PwLC symptoms, with some admitting they lacked confidence due to Long COVID being a new and largely unknown condition for which they had no training.

> *“I suppose the other challenge from my point of view is actually just setting up a new service from scratch, so you know that’s a big job in itself and setting up without the evidence base without kind of any benchmark you know to work with. And so, trying to do that alongside learning about a new disease you know with information coming out constantly.” [S4101]*

AHPs in integrated services also reported a lack of knowledge and confidence in working with people presenting with Long COVID due to the broad range of symptomatology.

> *“Hardest thing – so many different symptoms that you can have with Long COVID.” [S2204]*

Finally, less experienced AHPs in integrated services were reported to lack confidence in dealing with Long COVID cases as they saw fewer PwLC. This meant that in some areas increased pressure was placed on the limited capacity of senior staff.

A limitation within both integrated and dedicated rehabilitation services was the lack of medical input to both integrated and dedicated services and the challenges of onward referral to specialist secondary care.

> *“Having the right staff, having medical support in the team, that’s been a big gap I feel, and that’s still a gap and I think that’s really important given the complexity of patients coming through. We’re doing a lot of “safety netting” as AHP’s at the moment, but I don’t think, you know, that that’s our job, necessarily. We’ve had to take on some of that.” [S4101]*

## DISCUSSION

We explored the perceptions and experiences of community rehabilitation for Long COVID from the perspectives of patients, AHPs delivering community rehabilitation services, and NHS staff leading on the Long COVID response in four Scottish health boards. We were particularly interested in exploring the barriers and facilitators to Long COVID rehabilitation experienced by these three groups across the four different health boards. We found several barriers to accessing community rehabilitation for Long COVID across both dedicated and integrated services, albeit for different reasons, and not only during periods when non-essential services were restricted. We found that the uncertainties of Long COVID, including reluctance of GPs to diagnose, and the constantly evolving evidence-base were barriers to accessing rehabilitation services, and to AHPs confidently providing Long COVID rehabilitation. We also found that lack of or short-term funding, pre-COVID funding of community rehabilitation services, and reluctance to promote existing services to the Long COVID population were key operational barriers to Long COVID rehabilitation. Facilitators included flexible services with clear referral pathways, MDT working, sourcing professional information, and support from within and outwith health board areas. While patients may experience frustration around accessing services, and the evidence base for Long COVID rehabilitation is not firmly established, support and information received from an empathetic AHP was highly valued.

Barriers to accessing healthcare services for Long COVID have been reported in previous research focusing on GPs [12 24] and the views of people with Long COVID [25]. We have highlighted that specific barriers to accessing community rehabilitation for PwLC are principally related to unclear referral pathways and demand-capacity issues. Barriers to accessing the physical rehabilitation workforce are not a new phenomenon; Long COVID has arguably brought this previously ‘relatively neglected’ workforce into the spotlight [26] and highlighted the need for adequate human resourcing of rehabilitation services. This concurs with Baz et al’s [12] study of accessing healthcare support for Long COVID in England, which identified staff having to use existing, already stretched, services to support people with Long COVID. Brennan et al [24] reported that GPs (the principal gatekeepers to community rehabilitation services) found referral to community services challenging to negotiate. We identified a complex interplay of factors that impacted on referral from GP to community rehabilitation, including in some health boards a reluctance to promote services due to existing or anticipated capacity issues. Indeed, the dedicated Long COVID service in our study was quickly overwhelmed, demonstrating the real need for Long COVID rehabilitation, which the current epidemiology literature supports [3 27], and highlighting a capacity issue that needs to be urgently addressed.

There is currently no consensus on the optimal model of service delivery for Long COVID, beyond recommendations for it to be multilevel [15 18], multi-professional [15 28–30], person-centred [28], and adequately resourced [15]. Our findings support these recommendations. Services that are flexible to patients’ needs and offer a range of delivery modes were valued by patients and staff. The benefit of multidisciplinary team working was a key theme throughout our findings, from both staff and patients’ perspectives. However, the MDTs in our study were comprised of various AHPs and in some cases nurses and psychologists; access to medical specialists, as seen in some Long COVID dedicated MDT services or ‘one stop’ clinics [18] was notably lacking in the four participating health boards and identified as a limitation by participants.

The uncertainties around Long COVID, its diagnosis (particularly with the withdrawal of widespread testing), and optimal management have all been previously reported [31], and are inevitable given the newness of Long COVID as a condition and the as yet emerging evidence-base. It is encouraging that research on Long COVID rehabilitation is ongoing. It is vital to continue to explore Long COVID rehabilitation service delivery models, as well as the effectiveness of specific rehabilitation interventions, acknowledging that there is unlikely to be one that fits all contexts or geographical locations [15].

### Strengths and limitations

This is the first large scale study to explore community rehabilitation services for Long COVID across the Scottish context. We included the perspectives of those receiving and delivering services, as well as those responsible for their oversight. Data collection took place over two rounds, enabling us to capture some of the shifting practices inevitable with a new condition. Several researchers were involved at all stages of data analysis and interpretation, including people with lived experience of Long COVID. The study is not without limitations. We selected the four Scottish health boards based on geographical distinction and service delivery variance, as described in our 2019 national survey [14]. However, by the time we commenced data collection, approaches to Long COVID rehabilitation had already evolved meaning that there were fewer distinct service delivery differences between the four health boards. Nonetheless, our selected health board areas enabled us to investigate differences across the two principal modes of service delivery (dedicated and integrated) in Scotland at the time of writing. While we are confident we interviewed all key Long COVID leads in each participating health board and had high levels of AHP participation, patient recruitment was limited by low numbers of patients accessing Long COVID rehabilitation services.

### Implications for practice and research

There is likely to be an ongoing need for community rehabilitation for Long COVID, and no ‘one-size-fits-all’ model of service delivery. The findings presented here can be used to inform the integration of Long COVID into existing services, or to develop new dedicated rehabilitation services. Either way, service providers are required to provide adequate resource, clear referral and management pathways, and facilitate meaningful MDT working. This study represents the first stage in exploring community rehabilitation for Long COVID. We have also explored the perceptions of GPs and adults with Long COVID who have tried to access rehabilitation services (to be reported elsewhere). A realist evaluation of the services in the four health boards reported is ongoing. This evaluation will inform contextualised, evidence-based recommendations for Long COVID rehabilitation. The transferability of key findings from this Scottish-based study should be considered when considering their implications for different clinical and policy contexts.

## CONCLUSION

The organisational delivery of Long COVID community rehabilitation is complex and faces several challenges. We have provided a detailed understanding of the barriers and facilitators to Long COVID community rehabilitation from the perspectives of patients, AHPs, and Long COVID leads. The knowledge presented here can be used by those developing and delivering services for people with Long COVID. Community rehabilitation services have much to offer people with Long COVID, but requires adequate planning, publicity, and resource.

## Data Availability

All data produced in the present study are available upon reasonable request to the authors

## Acknowledgement

We are grateful to all our participants who gave their time to take part in interviews and to the local principal investigators from the health boards; Lynne Frew, Lynn Morrison and Gail Thomson-Patel, for their contributions to the LOCO-RISE study. We extend our sincere gratitude to the rehabilitation and administrative staff for supporting recruitment and data collection, and to Valery Burnett for her assistance with data collection.

## Author contributions

KC, ED, JM, LA, JP, JO, AL contributed to the study’s conception and design. TT, VB, JC, RM, JS undertook data collection. ED, KC, TT, JC, JS, ES, RM undertook analyses and drafted the first version of the manuscript. ED led future iterations of the manuscript. All authors read and commented on the manuscript and approved the final version of it. The corresponding author attests that all authors meet authorship criteria and that nobody meeting the criteria have been omitted. ED and KC have joint responsibility for the research conduct of the study, had access to the data, and controlled the decision to publish.

## Ethics approval

Ethical approval was granted from the Wales Research Ethics Committee 6 [21/WA/0118], and each of the four health boards granted R&D management approval.

